# Safety During Night Duty: Survey of 3885 Doctors Across India

**DOI:** 10.1101/2024.09.03.24312775

**Authors:** Rajeev Jayadevan, Deepa Augustine, TS Anithadevi, Reshmi Ramachandran, Joseph Benaven

## Abstract

**Background:** Reports of violence against doctors at the workplace are on the rise. In August 2024, a young female doctor was raped and murdered during night duty at her workplace in Kolkata, India. This incident prompted nationwide protests and a service shutdown by doctors’ organizations advocating for improved workplace safety.

**Aim:** This survey was undertaken by the Indian Medical Association to evaluate safety concerns during night shifts among doctors. With 3,885 individual responses, it is the largest study from India on this topic.

**Methods:** An online survey was sent to doctors across India through a Google form. There were 3885 responses within 24 hours. In addition to seeking suggestions to improve safety, three separate parameters were assessed:

1. 0-10 numeric rating scale of perception of safety
2. Availability of duty room
3. Access to bathroom while on night duty

**Results:**

1. Respondents were from several states. 85% were under 35 years. 61% were interns or postgraduate trainees. Women constituted 63%, aligning with the gender ratio in some MBBS courses.
2. Several doctors reported feeling unsafe (24.1%) or very unsafe (11.4%), totalling one-third of the respondents. The proportion of those feeling unsafe was higher among women.
3. A duty room was not available to 45% of respondents during night shifts.
4. Those with access to a duty room had greater sense of safety.
5. Duty rooms were often inadequate due to overcrowding, lack of privacy and missing locks, forcing doctors to find alternative rest areas.
6. One-third of available duty rooms did not have an attached bathroom.
7. In more than half the instances (53%), duty room was located far from the ward/casualty area.
8. Suggestions to enhance safety included increasing the number of trained security personnel, installing CCTV cameras, ensuring proper lighting, implementing the Central Protection Act (CPA), restricting bystander numbers, installing alarm systems, and providing basic amenities such as secure duty rooms with locks. For detailed information, refer to Table 13, the synopsis and the verbatim comments section.

**Conclusions:** Doctors across the country, particularly women, report feeling unsafe during night shifts. There is substantial scope for improving security personnel and equipment in healthcare settings. Modifications to infrastructure are essential to ensure safe, clean, and accessible duty rooms, bathrooms, food, and drinking water. Adequate staffing, effective triaging, and crowd control in patient care areas are also necessary to ensure that doctors can provide the required attention to each patient without feeling threatened by their work environment.

## 1. Introduction

### Background

Healthcare professionals are required to take night shifts as part of their training during internships and postgraduate studies, and they continue to work night duties in both private and public sectors throughout their careers. This aspect of the profession makes them vulnerable to various forms of workplace violence.

Public anger is frequently directed at doctors, who are perceived as the leaders in a healthcare team. In crowded settings like outpatient facilities, emergency rooms, or ICU waiting areas, doctors on the frontline are particularly vulnerable to sudden and unprovoked attacks. Violence can be triggered by perceived or real deficiencies in healthcare delivery, adverse outcomes, payment disputes, and substance abuse. While most instances of violence are spontaneous, some cases, such as the recent incident in Kolkata, are premeditated. These acts often result in more severe and sometimes fatal outcomes due to the advanced planning involved and the backgrounds of the perpetrators.

Although targeted at doctors, workplace violence ultimately impacts the quality of care provided to the public. For example, doctors may become hesitant to undertake potentially risky life-saving procedures and may prefer to refer patients to other centres. Night shifts increase doctors’ vulnerability to violence due to reduced staff, the cover of darkness, and the presence of individuals under the influence of alcohol or drugs. Factors such as the lack of dedicated and secure duty rooms, their distance from the workplace, and the need to walk a significant distance to access facilities further heighten their risk. Women doctors face greater risks in these situations.

Globally, workplace violence is a known issue. A 2017 study by the Indian Medical Association found that over 75% of doctors in the country have experienced workplace violence, while 62.8% are unable to see their patients without any fear of violence [1,2]. Another study reported that 69.5% of resident doctors encounter violence while at work [3]. Exposure to violence is known to lead to fear, anxiety, depression, and post-traumatic stress disorder among doctors [4].

Healthcare related violence is a worldwide problem. In China, a survey among medical staff in children’s hospitals revealed that 68.6% had suffered verbal or physical violence in the past year [5].

Perceptions of safety significantly influence job satisfaction among doctors [6]. The recent rape and murder of a young woman doctor in Kolkata India triggered widespread protests among the medical fraternity [7]

This survey was conducted to address the following research question: “How do doctors in India perceive their safety during night shifts? What environmental factors influence their sense of safety, and what suggestions do they have for improving their safety?”

## 2. Objective

1. To assess three safety parameters pertaining to night duty
  a. 0-10 scale of perception of safety
  b. availability of duty room
  c. Access to bathroom while on night duty
2. To collect suggestions from individual doctors about how their personal safety can be improved.

## 3. Methodology

### Study Design

An online survey of 10 questions was sent to doctors across India through Google form. There were 3885 responses within 24 hours. The survey was cross-sectional and anonymous, which was to encourage individual doctors to provide their frank opinion. Data was collected through a Google form on 17 August 2024, and the results were accessed on a Microsoft Excel spreadsheet.

### Parameters Assessed

1. Perceived Safety Participants chose a number on a numeric rating scale of 0-10 indicating their sense of safety, with 0 being the worst and 10 being the best. A score of 0-3 was classified as unsafe, while 8-10 was interpreted as safe. 0 was called very unsafe, while 10 was termed safest. Those who scored 4-7 were classified as “uncertain”
2. Duty Room Availability: Yes/No question about whether they have a duty room.
3. Bathroom Accessibility: Yes/No question about whether the duty room has an attached bathroom. Distance to bathroom: to see if it was nearby or otherwise.
4. Suggestions from individual doctors about how their personal safety can be improved.

## 4. Statistical Analysis

Descriptive statistics were employed to evaluate the baseline characteristics of the survey population, using counts and percentages for qualitative variables, with bar charts and pie charts for visual representation. To assess the association between safety perceptions and other variables, chi-square tests or Fisher’s exact tests were applied, depending on cell counts. The analysis was conducted with a 95% confidence interval, setting the significance level at 5%. Data were entered into Microsoft Excel and analysed using SPSS version 20.0.

## 5. Results

The survey conducted by IMA Kerala State Branch in August 2024, captures data from 3,885 respondents from various states across India, providing insights into gender, age, designation, duty room availability, and safety perceptions during night duty.

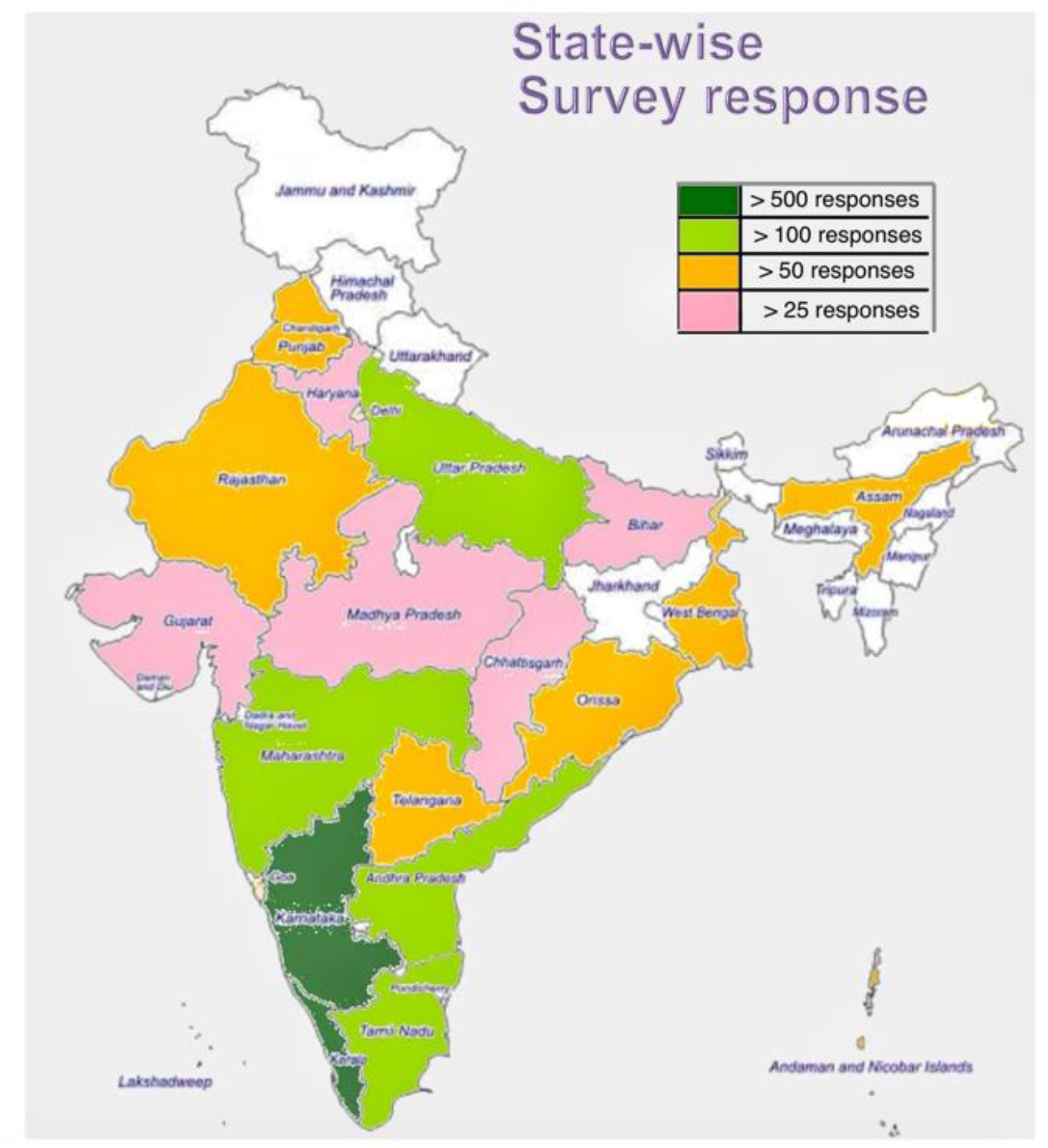

The survey received 3885 responses from doctors across India. The states are colour-coded according to the number of responses received. See table 10 for details.

**Graph 1:**
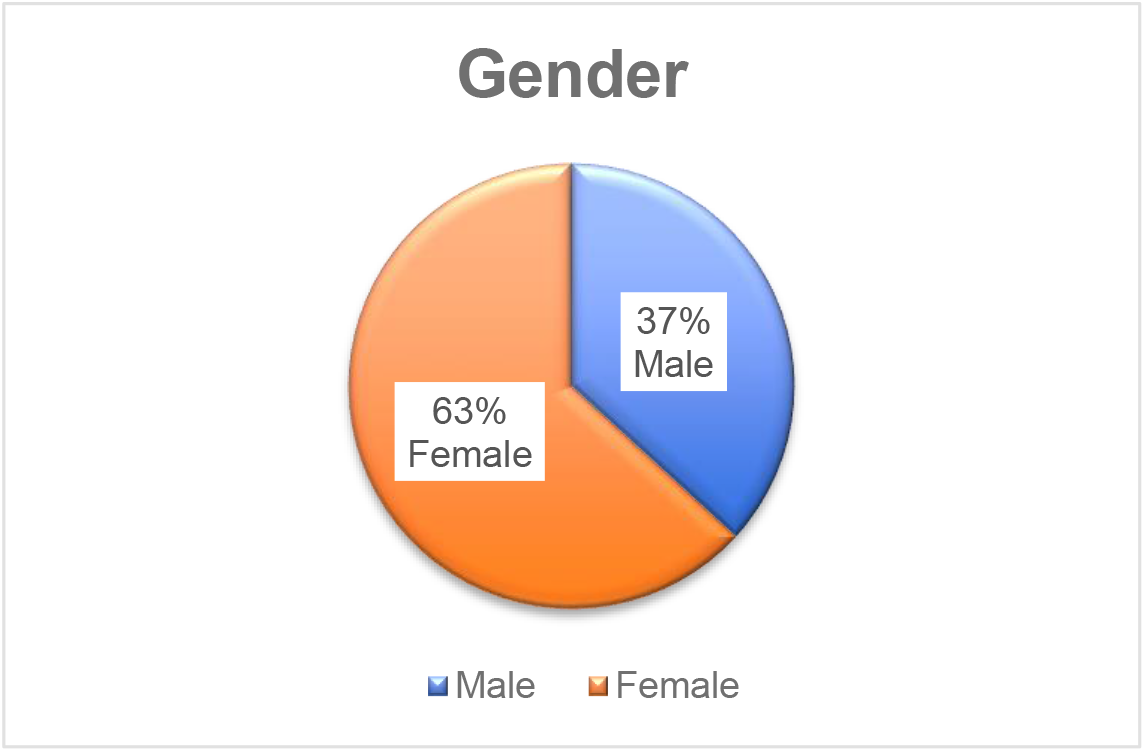
The majority of the 3,885 respondents, were women, (63%) reflecting current gender distribution trends among MBBS students in certain states such as Kerala [8].

**Graph 2.**
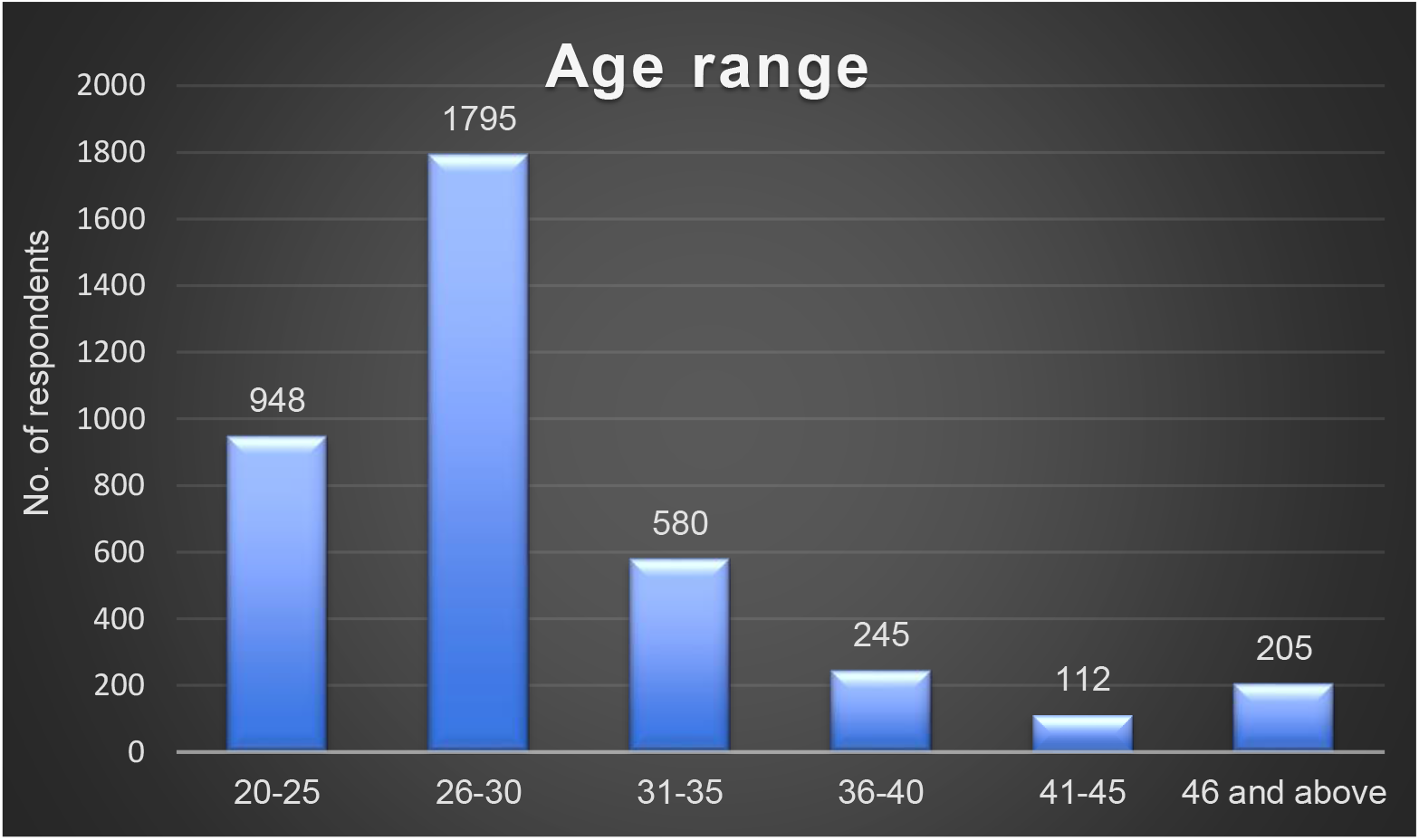
Age distribution of survey respondents. 85% of the respondents were at or below age 35. Two-thirds were aged 30 or below. The majority (46.2%) fall within the 26-30 year age range, with 24.4% in the 20-25 year group. Smaller proportions are seen in older age categories, with just 5.3% aged 46 years and above.

**Graph 3:**
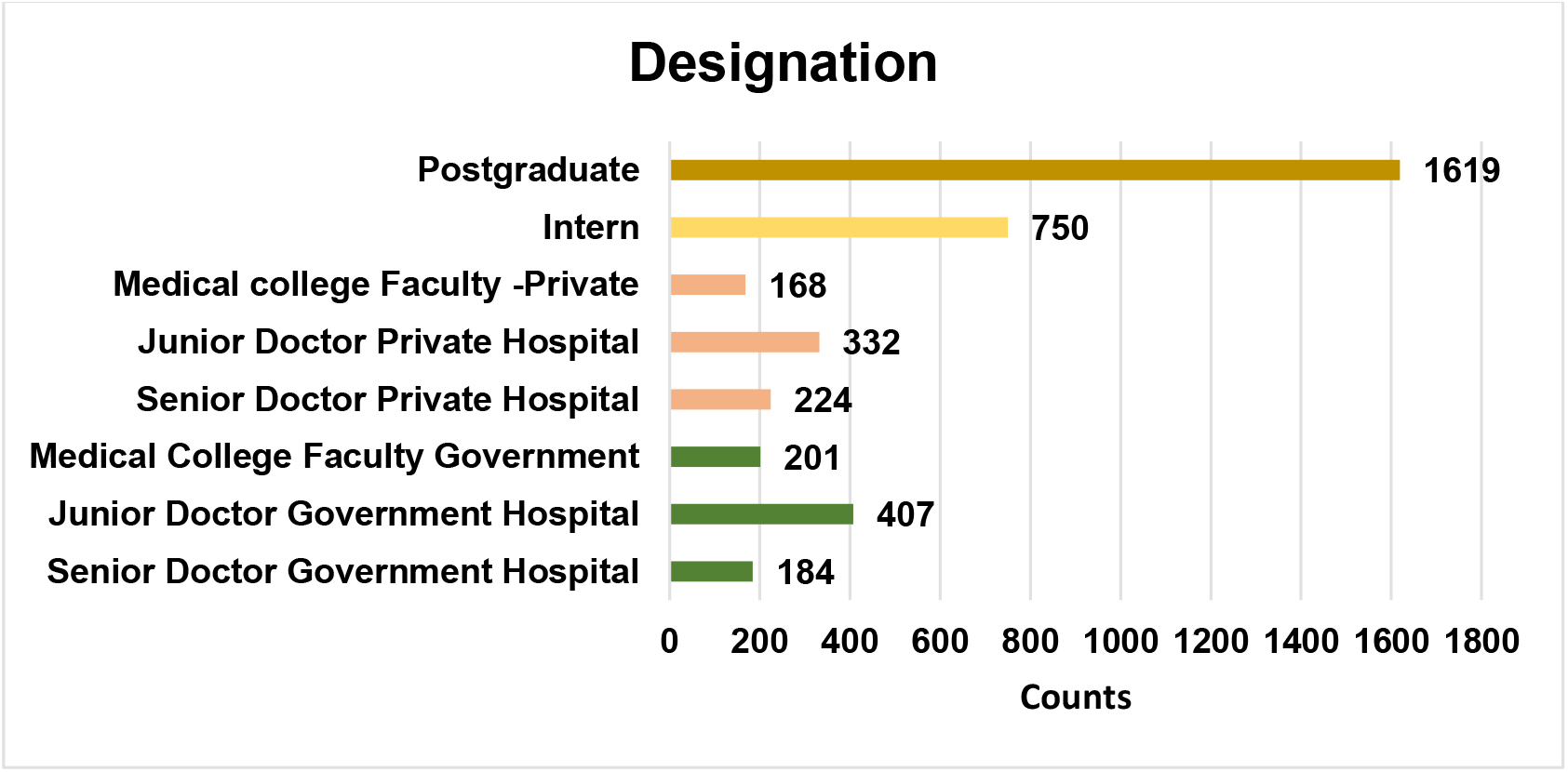
Designation of doctors who took the survey.

**Graph 4:**
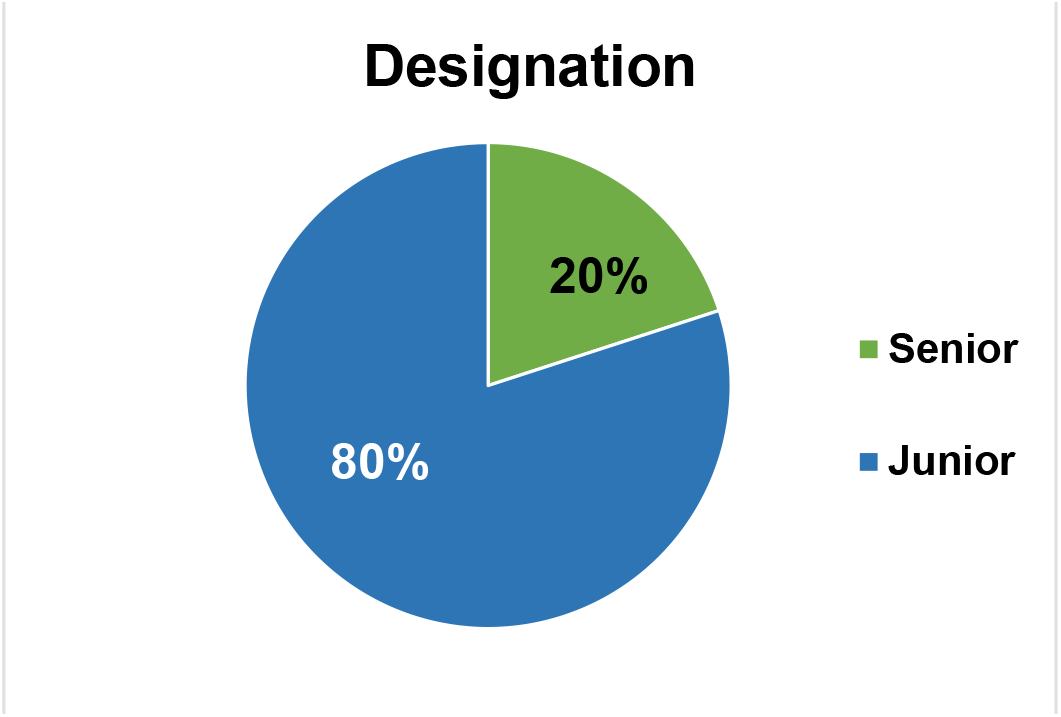
Professional seniority status. The designation distribution among study participants shows that the largest group consists of postgraduates (41.7%), followed by interns (19.3%), with a combined representation of 61%.

Junior doctors in government hospitals make up 10.5%, while their counterparts in private hospitals account for 8.5%. Senior doctors and medical college faculty were evenly distributed, with senior doctors from private hospitals at 5.8%, government hospitals at 4.7%, and medical college faculty at 5.2% in government and 4.3% in private institutions. The distribution reveals that a majority of participants (80%) are junior doctors. This indicates that the survey primarily captured the perspectives of those in relatively junior positions (Graph 5).

**Graph 5:**
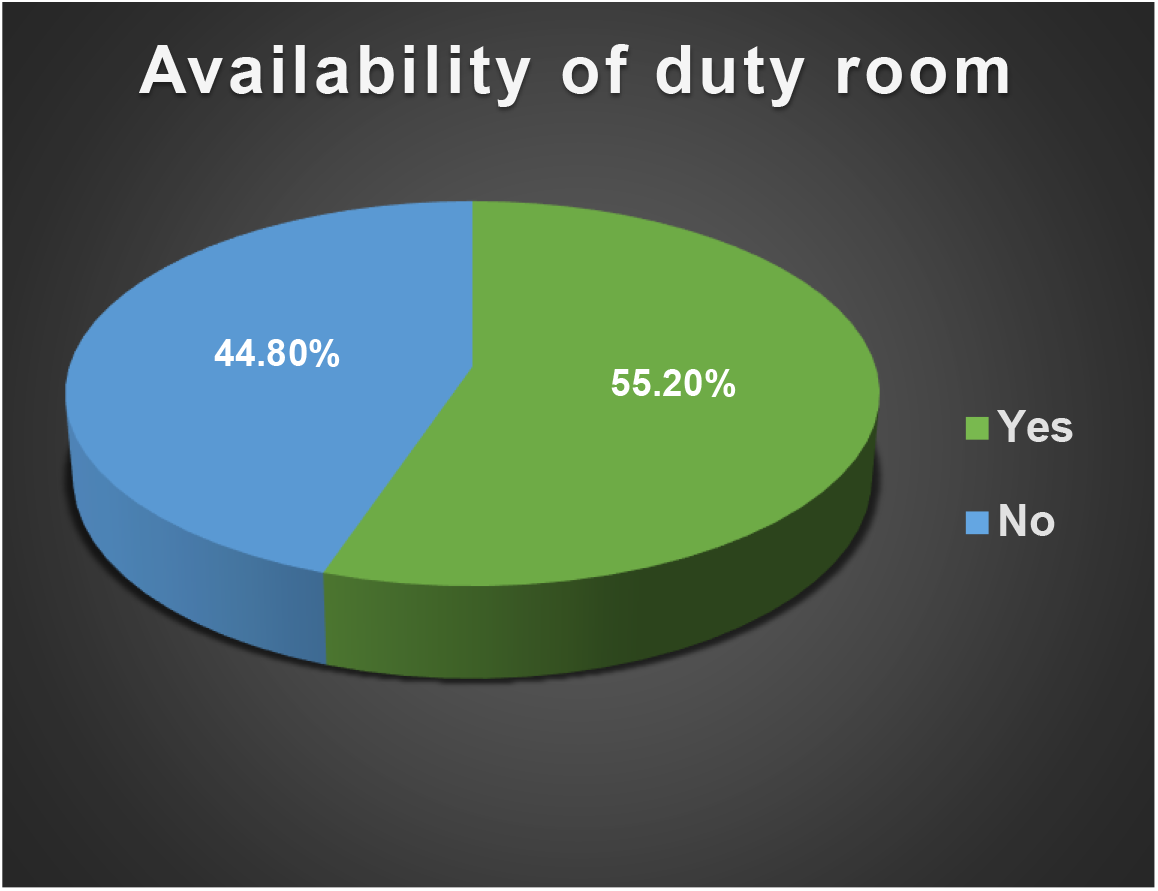
Duty room availability. The assessment of duty room availability among the participants shows that 55.2% have access to a duty room, while 44.8% do not.

**Graph 6:**
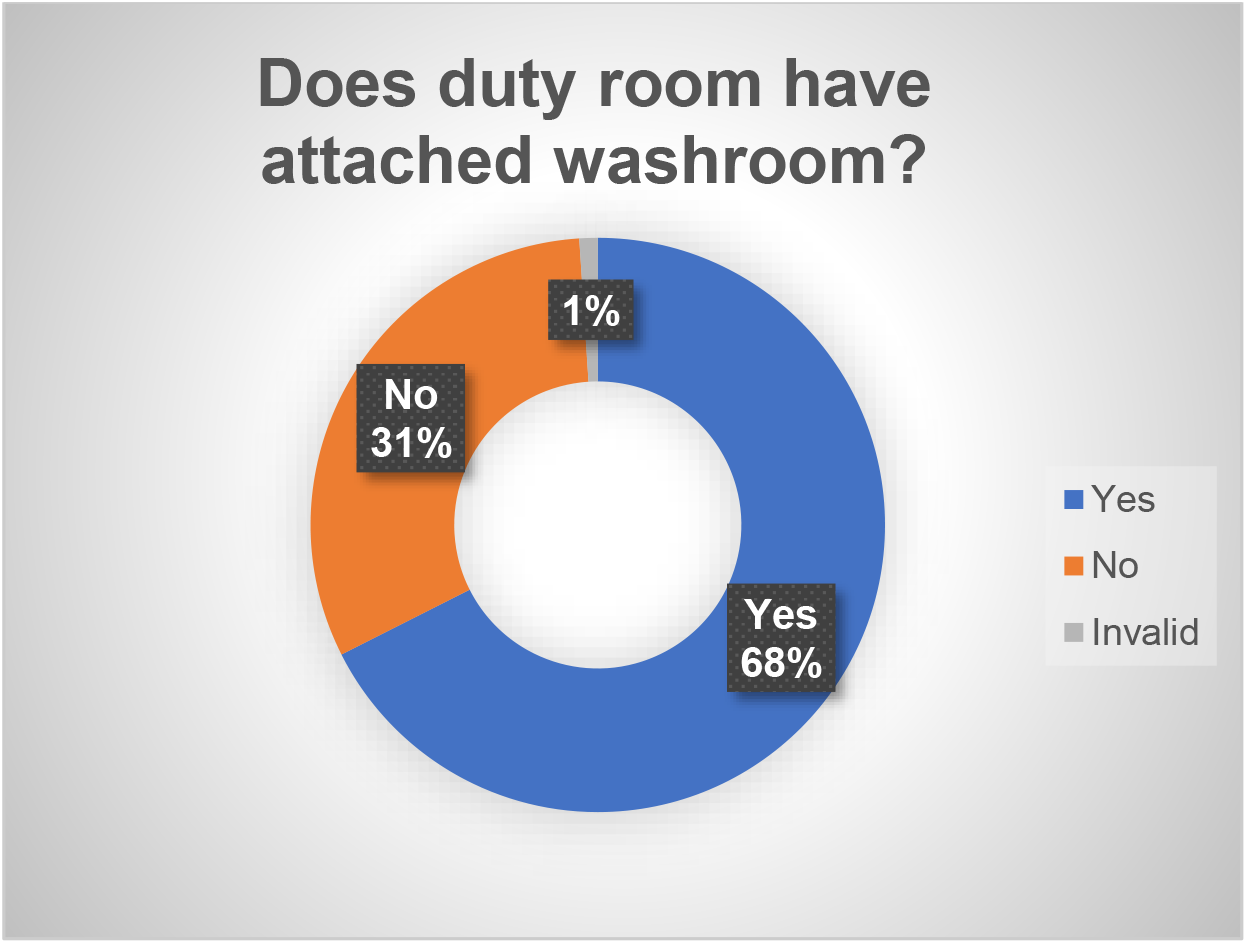
Duty Room with attached washroom/restroom. Among the 2,145 participants with access to a duty room, 67.6% reported having a duty room with an attached washroom/restroom, while 31.4% did not. A small percentage (1.03%) gave invalid responses.

**Graph 7:**
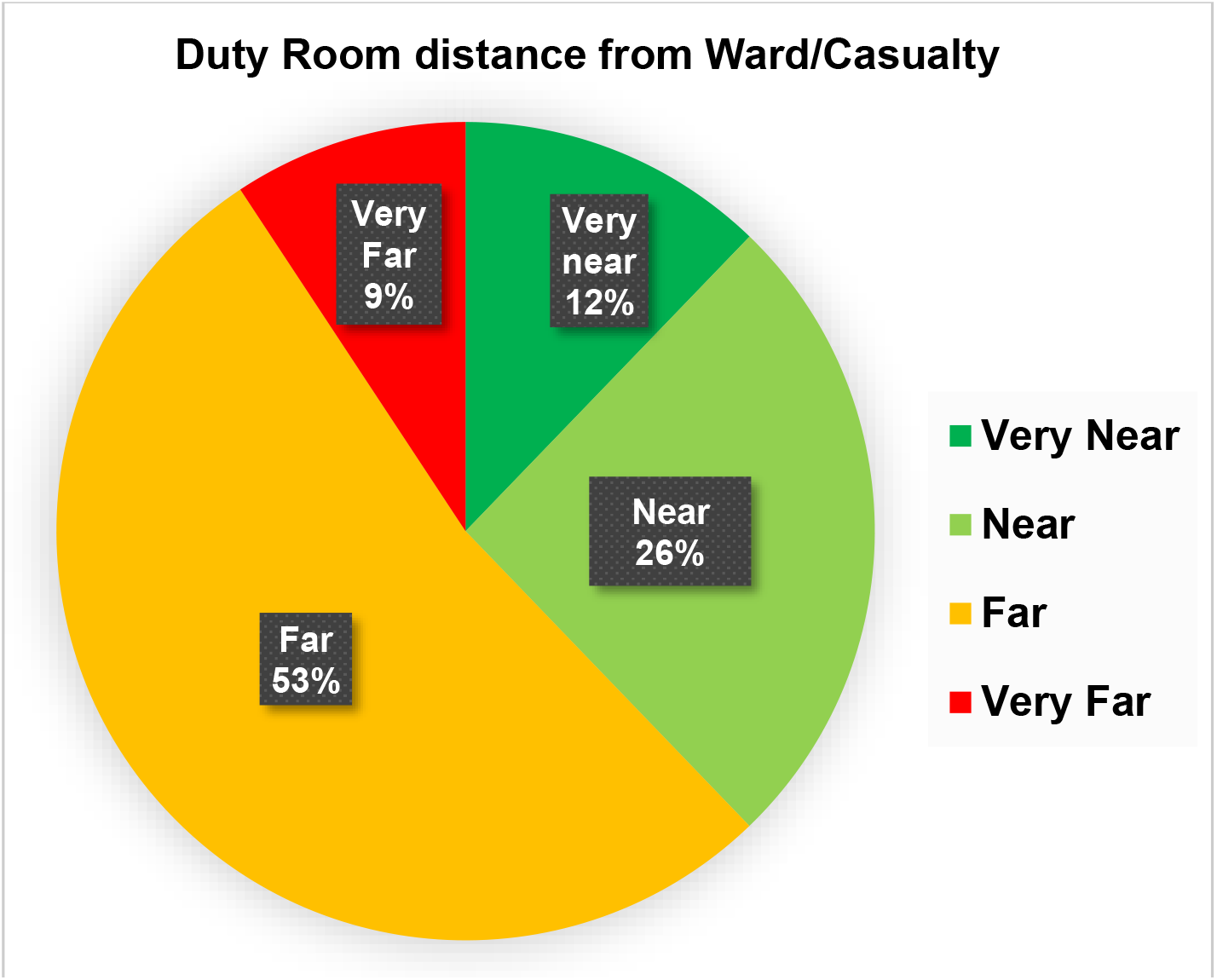
Distance of duty room from ward/casualty. Among the participants with access to a duty room, 52.9% reported that their duty room was located far away from the ward or casualty area (100-1000 meters). A majority of doctors must walk a significant distance from their duty rooms to reach the ward or casualty area, which can pose a safety risk at night if the path is not well-lit and secure.

**Graph 8:**
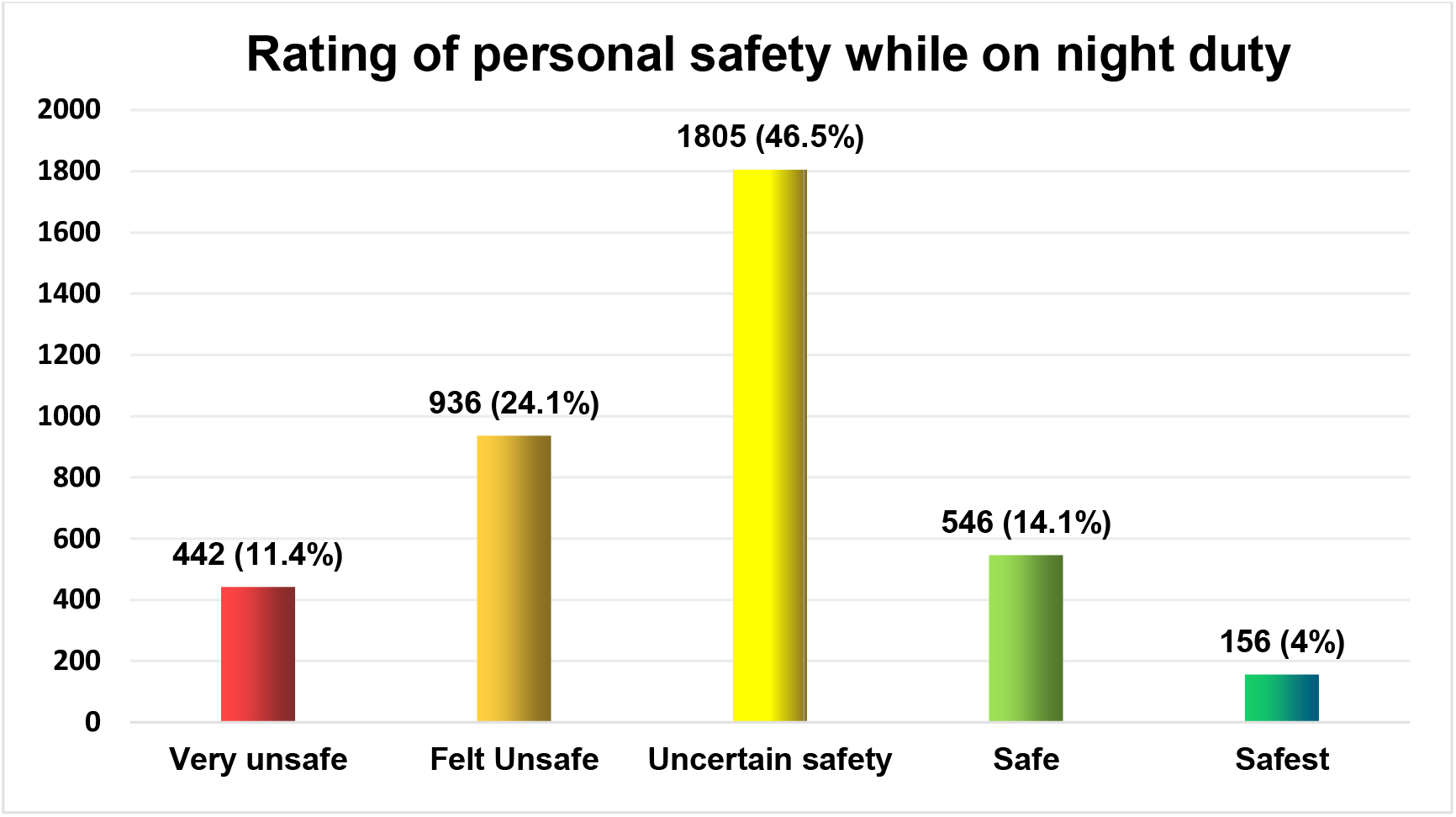
Self-Reported Safety Level in workplace. The subjective assessment of safety during duty hours shows that a significant portion, 24.1%, felt unsafe, and 11.4% considered their situation very unsafe, with a total of 35.5% feeling unsafe to various degrees. On the other hand, 14.1% felt safe and 4% felt it was the safest. 46.5% of respondents reported “Uncertain Safety”, reflecting mixed feelings or uncertainty about their safety.

Women report higher levels of feeling unsafe or very unsafe (36.7%) compared to men (32.5%), with this finding being statistically significant (p < .0001). Age also plays a role, with younger respondents (20-25 years) feeling less safe compared to older groups, and this age-based difference is significant (p < .0001). Regarding designation, juniors generally feel less safe compared to seniors, with this discrepancy also being significant (p < .0001).

**Graph 9:**
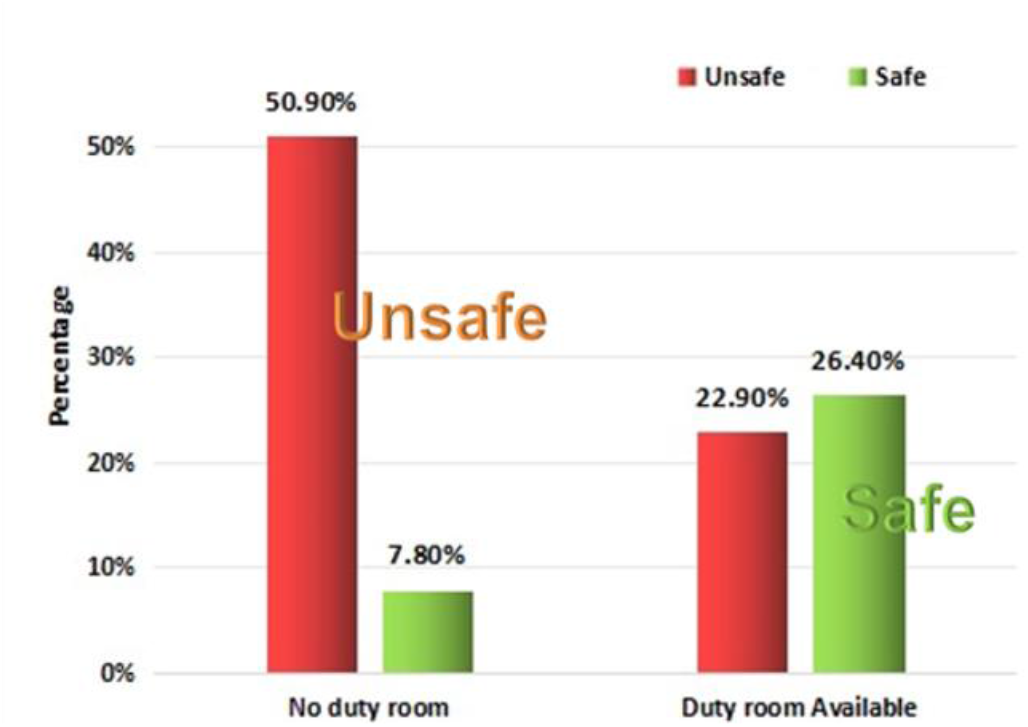
Duty room and sense of safety. Association between duty room and sense of safety: Two parameters relating to safety perception are displayed here. The red bar is the proportion of those who felt unsafe, scoring 0-3 on a numeric rating scale of 0-10, while the green bar represents those who felt safe, with a score of 8-10.

When duty room was not available, more doctors felt unsafe (tall red bar) and fewer doctors felt safe (short green bar).

This shows that availability of a duty room significantly impacts safety perceptions, with those without access feeling unsafe (50.9%) compared to those with a duty room (22.9%) (p < .0001). Similarly, 26.4% of those with a duty room reported feeling safe compared to only 7.8% without.

Likewise, the presence of a duty room with an attached washroom is associated with better safety perceptions; more of those without such facilities feel unsafe (44.8%) compared to those with the facility (20.2%) (p < .0001). Similarly, more of those with an attached washroom felt safe (30%) compared to only 10.8% without. This shows a clear correlation between the availability of basic facilities and safety perception.

Government healthcare workers reported higher levels of insecurity, with 17.05% feeling “Very Unsafe” and 27.4% feeling “Unsafe”, totalling 44.5%. In contrast, only 5.52% and 12.02% of private healthcare workers reported feeling “Very Unsafe” and “Unsafe,” respectively, totalling 17.5%. The classification was described earlier, in methodology.

Similarly, a larger percentage of private healthcare workers felt “Safe” (28.04%) compared to those in government institutions (8.71%). Additionally, 10.22% of private healthcare workers felt the “Safest,” whereas only 1.89% of government workers shared this level of confidence in their safety.

Adding up, 38.3% felt safe in the private sector, while only 10.6% shared the same sense of safety in the government sector.

This section of analysis did not include interns or postgraduate trainee doctors, as they were not subclassified into public or private sector in the survey.

## Suggestions to improve safety

Numerous suggestions were raised by 3885 respondents. Some individuals contributed several, while others mentioned one or two key points. All of these were individually read by the study authors in repeated sittings, categorised according to the theme and quantified by frequency. All of this qualitative analysis was done manually using pencil and paper. The reasons for doing this manually was to acquire a deeper understanding as well as to eliminate any misreading due to language limitation. Subsequently, a statistical analysis was performed.

The following are suggestions ranked according to frequency of keyword use. Those which were mentioned the most are given on top. Details are given in the subsequent section.

### Synopsis of suggestions

#### The following synopsis summarizes the numerous comments in each category. A verbatim comments section is provided further below, offering an unfiltered view of the concerns raised

##### Security

Inadequate security at hospital premises was reported by a large number of doctors. Hospitals often employ low-wage security personnel to reduce costs. Many commented that the available security personnel were weak and frail individuals who themselves seemed to need protection and were the first to run away at the first sign of trouble. Doctors have expressed a preference for exservicemen as security guards, and able-bodied men in their 30’s to 40’s as bouncers in high-risk areas such as casualty and ICU lobby where skirmishes are common. Some women doctors indicated the need for female security personnel. Having a police checkpost is ideal. Security is particularly lacking in smaller peripheral hospitals where doctors may be alone with limited staff. Security personnel must be proportionate to the size of the hospital and must conduct regular rounds of the premises. Doctors who need to walk to different areas of the hospital during late hours—such as the blood bank, scan room, ward, lab, duty room or parking lot—should be accompanied by a security escort.

1. **CCTV camera:** Functioning cameras are required in multiple areas of healthcare establishments, and they must be manned by trained personnel who have the resources to take action should a potential problem be detected.
2. **CPA (National Law that protects healthcare workers)** Doctors were emphatic about the need for a CPA or Central Protection Act that enables them to work safely in any healthcare establishment. The absence of such a law encourages unscrupulous behaviour. It is equally important to file an FIR on time and to enforce the law when violence occurs. Awareness of such a law will deter violence. The presence of a grievance redressal forum at each facility will prevent minor discontent from escalating into violence.
3. **Duty room:** Inadequate facilities were widely reported. When a doctor takes a night call, they need a place where they can rest for some time. The duty room must be secure and clean, with bolts on the door and an accessible, secure bathroom. Many suggested that women and men have different duty rooms and bathrooms. Interns as well as postgraduates must have duty rooms, and these must not be shared by other hospital staff. The room must be located away from where bystanders are waiting, but not in dark, isolated and deserted parts of the building. Currently, in the absence of duty rooms, many doctors are forced to go to insecure places like a seminar room or an empty cot on a ward or OPD to lie down, which occasionally leads to experiences such as theft and even assault.
4. **Lighting** is required throughout the areas where doctors and staff are expected to be using while doing night calls. Doctors reported several instances of dark unlit corridors, duty rooms located in isolated parts or floors of the building and parking lots without security or lights.
5. **Restricting the number of bystanders** is crucial in healthcare settings. Doctors reported instances of being surrounded by crowds while attending to road accident victims or performing procedures in the casualty area. This is both unnecessary and dangerous, as some individuals may be under the influence of alcohol or drugs, increasing the risk of violence due to ignorance or aggression. Ideally, only female bystanders should be permitted in wards at night. All building entrances must be secure and monitored to prevent unauthorized access to patient care areas or isolated sections of the facility. Construction workers should not be present on the premises at night. Bystanders should be given wristbands for identification and should not be allowed entry without them.
6. **The absence of locks and bolts on duty rooms** has been highlighted by many doctors, and this is a problem that can be fixed with minimal investment. Doctors said that they often do not know who is knocking on their duty room door until they open it, this poses a significant security risk. Some duty rooms do not have complete walls, a few others only had tinted glass partitions that did not provide adequate privacy.
7. **Panic Button:** Despite substantial investment in infrastructure and personnel, violence can occur at any time in healthcare settings. In such situations, there must be a panic button, emergency helpline, or specialized alarm system to summon immediate assistance. Known as code grey or code white in various facilities, this practice is already implemented in several tertiary care centers and should be made universal.
8. **Alcohol and drugs** play a major role in healthcare violence. Bystanders who are inebriated should not be allowed to enter casualty, and this requires protocols and manpower in the security/triage department.
9. **Food and Water:** Food and water are fundamental needs for doctors on night shifts. Providing these essentials near their work areas eliminates the need for doctors to walk through dark and unsecured parts of the campus to obtain them. Additionally, vending machines for sanitary napkins should be installed, and designated rooms should be provided for doctors who are nursing mothers.
10. **Grievance Redressal or Helpdesk:** A help desk should be established outside casualty areas or wards where bystanders can direct their questions, rather than interrupting doctors. This also protects doctors from becoming convenient targets for even minor grievances. Additionally, having trained staff available for counselling can reduce some of the doctors’ workload in crowded settings.
11. **The support of administration** is essential to promote a safe and secure work environment. Interns and postgraduates highlighted the need to reduce the toxicity at the workplace, emphasising their right to be treated fairly. Periodic audits and appraisal meetings of workplace safety will help reduce the sense of fear that doctors feel while taking night calls.
12. **Animal attacks** were mentioned by several doctors. Stray dogs and snakes were the most common. A well-maintained, walled, clean campus will eliminate such threats.
13. **Non-clinical night duty:** Some trainee doctors questioned the necessity of performing night duties in non-clinical departments such as pathology and microbiology.

## A sample of comments verbatim: they provide insight into individual doctors’ concerns

1. “I have felt bad touches many times working in crowded casualty”
2. “The duty rooms were at the end of a deserted corridor without much light, always felt unsafe and always carried a pepper spray and foldable knife in my bag.”
3. “I don’t feel safe.”
4. “Security personnel run away when any situation arises.”
5. “Only one bystander to be allowed per patient.”
6. “Anyone can knock on the duty room door. We open without knowing who the person outside is.”
7. “Providing security alone is not enough, need law which punishes culprits.”
8. “Implement CPA” (Central Protection Act)
9. “If ESMA applies to us, protection too should.” (Essential Services Maintenance Act)
10. “I am concerned about my female colleagues.”
11. “We need an airport-like safe zone.”
12. “During 70% of my duty days, I had to handle drunk mobs at night.”
13. “10 people (bystanders) stand around me when I take RTA (road traffic accident) patient’s history.”
14. “Institutions must have written policy on safety at workplace.”
15. “Doctors have to do ward boy work due to lack of labour force.”
16. “Not just tertiary care hospitals/medical colleges, doctors are working at peripheral hospitals as a part of bond service or training or DRP, so, security and safety should be provided at every work place”
17. “Security is an old, short, skinny man most of the times.”
18. “We have CCTV cameras everywhere and security personnel too, so no issues.”
19. “I do need a place to rest, somewhere with a lock and a place to use as a restroom.”
20. “Training of postgraduates to be non-toxic.”
21. “We are just provided some unused ward for all female duty doctors with one untidy washroom, and that ward is barely cleaned.”
22. “We often get attacked by patient bystanders in casualty and ICU.”
23. “We have faced atrocities of patient bystanders.”
24. “Provide a gun to all the doctors, and guards of younger age.”
25. “Security Jo Jagada Hone par sabse Pehle nahin Bhaage.” (We need security who do not run away at the first sign of trouble.)
26. “I have faced serious security threats as a woman doing night duties.”
27. “If we need to use the washroom, we need to go to hostels in the middle of the night when it is pitch dark, no security.”
28. “Postgraduates sleep in OPD as no room available.”
29. “I have faced exhibitionism from a middle-aged man in an outpatient department.”
30. “It is scary to run to different wards for code blue at night.”
31. “Approval for use of Self Defence tool for healthcare workers.”
32. “If we scream, no one will hear.”
33. “OPD’s should have a double door system. One for entry and one for escaping violent attacks from patients or bystanders.”
34. “Authorities don’t take our complaints seriously, they act as if this is how universally things are.”
35. “Do not make us go to deserted places and corridors at night.”
36. “Pls give a room with locks”
37. “A wearable panic button for every doctor on call”
38. “As I am working in a private setup, things are better”
39. “Just one duty room provided for all female PG’s and interns which will eventually become crowded, and we search for some other places to take rest which is unsafe”
40. “Though we don’t have a great duty room, we feel pretty safe, mainly because our seniors are open to our concerns and show genuine interest when we raise an issue. We also have good security guards.”
41. “Periodic safety audit to be done”
42. “…we are human beings, can’t work 36-48 hours without rest”
43. “Let security staff accompany the doctor at odd hours”
44. “Security needed in all ward/ICU/red zone, wherever doctor need to disclose death”
45. “Security guards looks not so strong.”
46. “Avoid more than 24 hours duty….one will be very weak physically and mentally to overcome any stressful situations”
47. “Most unsafe are the rural postings”
48. “I work in emergency medicine. I feel safe”
49. “One patient, one relative policy”
50. “Security to patrol isolated corridors. Lifts to have 24/7 security”
51. “Department is cool and safe, but rural posting hard to imagine”

## 6. Discussion

This is the largest study on healthcare violence done among doctors in India. Over one-third of the doctors reported feeling unsafe while doing night duty. More than one out of ten felt totally unsafe, giving the lowest score of zero out of ten for sense of safety. The feeling of being unsafe was worse among women, and among younger doctors. Doctors of age 20-30 years had the lowest sense of safety (p < 0.001, Table 9), this group largely consists of interns and postgraduates.

**Table 1:** Gender distribution.

| Gender | Frequency | Percentage |
| --- | --- | --- |
| Male | 1425 | 36.7% |
| Female | 2460 | 63.3% |
| Total | 3885 | 100% |

**Table 2:** Age distribution.

| Age | Frequency | Percentage |
| --- | --- | --- |
| 20-25 | 948 | 24.4% |
| 26-30 | 1795 | 46.2% |
| 31-35 | 580 | 14.9% |
| 36-40 | 245 | 6.3% |
| 41-45 | 112 | 2.9% |
| 46 and above | 205 | 5.3% |
| **Total** | **3885** | **100%** |

**Table 3:**
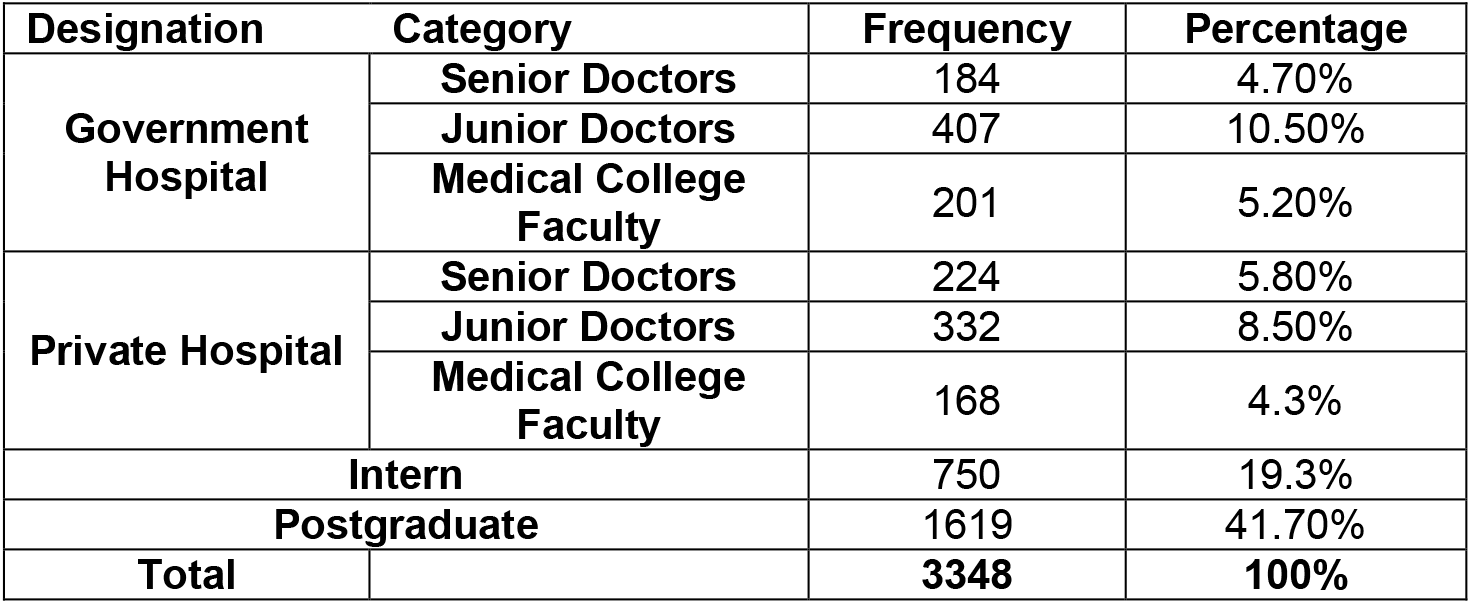
Distribution of designation of the doctors.

**Table 5:** Assessment of duty room availability.

| Availability of duty room | Frequency | Percentage |
| --- | --- | --- |
| Yes | 2145 | 55.20% |
| No | 1740 | 44.80% |
| Total | 3885 | 100% |

**Table 6:** Duty room with attached washroom facility.

| Duty Room with Attached washroom | Frequency | Percentage |
| --- | --- | --- |
| Yes | 1449 | 67.55% |
| No | 674 | 31.42% |
| Invalid | 22 | 1.03% |
| Total | 2145 | 100% |

**Table 7:** Distance of duty room from Ward/Casualty.

| Distance from Duty Room and Ward/Casualty | Frequency | Percentage |
| --- | --- | --- |
| Very Near | 262(<50 metres) | 12.21% |
| Near | 549(50-100 metres) | 25.59% |
| Far | 1135(100-1000 metres) | 52.92% |
| Very Far | 199 (>1000 metres) | 9.28% |
| Total | 2145 | 100% |

**Table 8:**
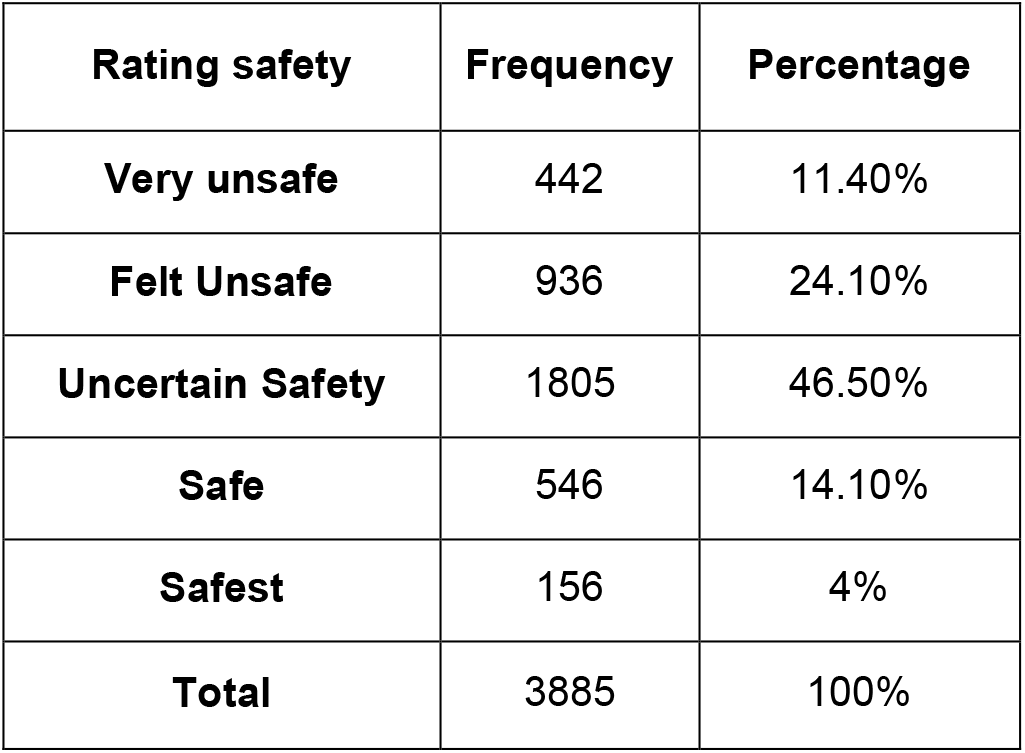
Subjective assessment of safety during duty hours. Participants chose a number on a numeric rating scale of 0-10 indicating their sense of safety, with 0 being the worst and 10 being the best. A score of 0-3 was classified as unsafe, while 8-10 was interpreted as safe. 0 was called very unsafe, while 10 was termed safest. Those who scored 4-7 were classified as “uncertain”

| Rating safety | Frequency | Percentage |
| --- | --- | --- |
| Very unsafe | 442 | 11.40% |
| Felt Unsafe | 936 | 24.10% |
| Uncertain Safety | 1805 | 46.50% |
| Safe | 546 | 14.10% |
| Safest | 156 | 4% |
| Total | 3885 | 100% |

**Table 9:** Safety perceptions during duty hours by demographic and facility parameters. The data shows significant variation in safety perceptions during duty across different demographics and environmental factors.

| Parameter |  | Very Unsafe | Felt Unsafe | Uncertain in safety | Safe | Safest | Significance |
| --- | --- | --- | --- | --- | --- | --- | --- |
| Gender | Male | 12.00% | 21.30% | 44.40% | 16.60% | 5.60% | Significant (p < .0001) |
|  | Female | 11.00% | 25.70% | 47.60% | 12.60% | 3.10% |  |
| Age | 20-25 Years | 11.30% | 29.30% | 45.10% | 11.60% | 2.60% | Significant (p < .0001) |
|  | 26-30 Years | 10.50% | 24.20% | 49.90% | 12.40% | 3.10% |  |
|  | 31-35 Years | 12.90% | 21.70% | 43.30% | 16.90% | 5.20% |  |
|  | 36-40 Years | 18.40% | 16.70% | 42.40% | 16.70% | 5.70% |  |
|  | 41-45 Years | 10.70% | 17.90% | 44.60% | 20.50% | 6.20% |  |
|  | 46 and above | 7.30% | 18.00% | 37.10% | 25.40% | 12.20% |  |
| Designation | Senior | 11.70% | 16.30% | 43.80% | 21.10% | 7.10% | Significant (p < .0001) |
|  | Junior | 11.30% | 26.00% | 47.10% | 12.30% | 3.20% |  |
| Duty Room Available | Yes | 5.60% | 17.30% | 50.70% | 20.20% | 6.20% | Significant (p < .0001) |
|  | No | 18.40% | 32.50% | 41.30% | 6.50% | 1.30% |  |
| **Duty Room with Attached Wash Room Facility** | **Yes** | 5.60% | 14.60% | 49.80% | 22.70% | 7.30% | Significant<br>(p < .0001) |
|  | **No** | 14.90% | 29.90% | 44.40% | 8.80% | 2.00% |  |

Chi square test, $p < 0.0001$ shows significance

**Table 10:**
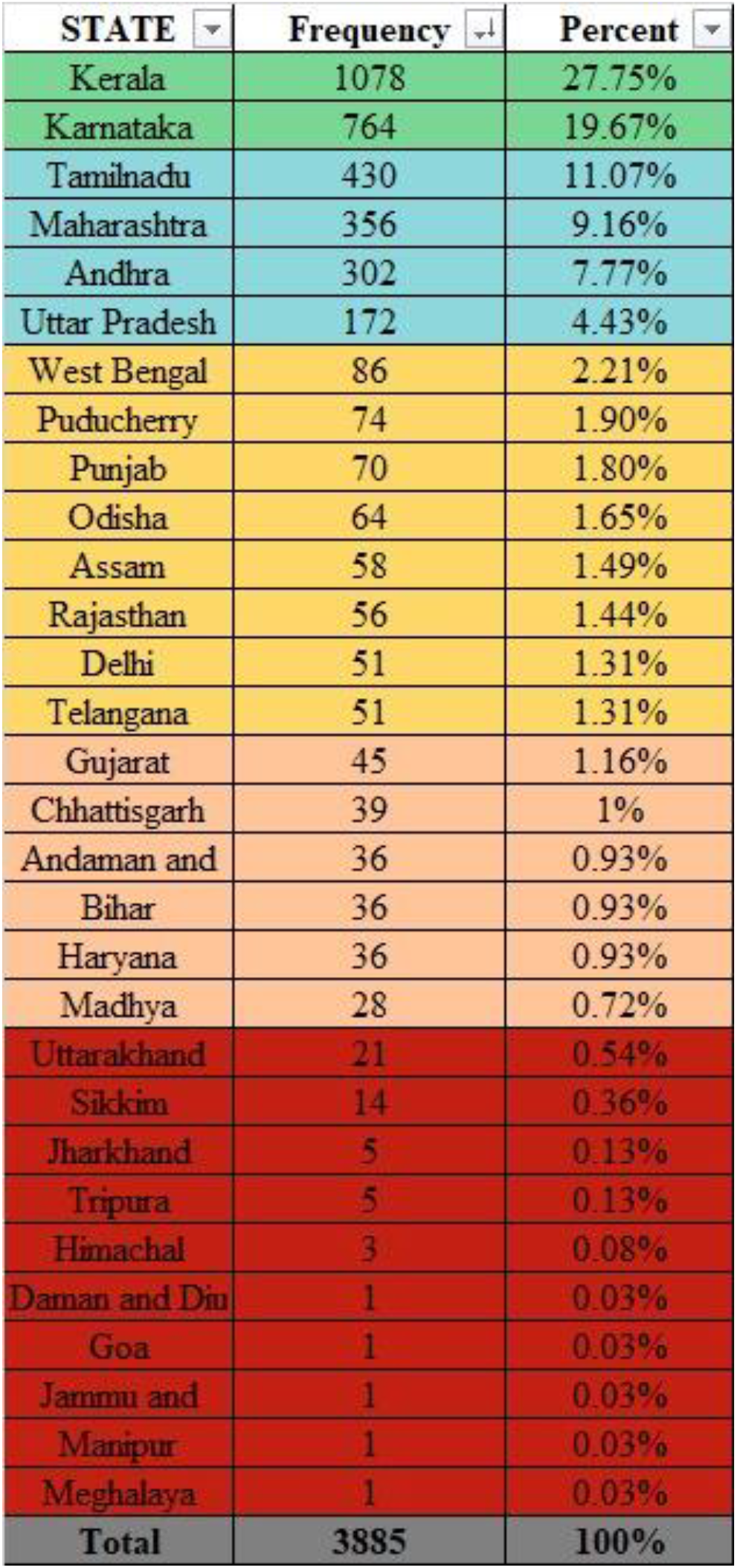
Statewise distribution of responses. The distribution of statewide responses reveals that the largest representation comes from Kerala (27.7%) and Karnataka (21.1%). More than a hundred individual responses each came from U.P (4.4%), Andhra Pradesh (7.2%) Maharashtra (9.2%), and Tamil Nadu (11.1%). More than 50 responses each were obtained from Assam, Delhi, Odisha, Puducherry, Punjab, Telangana and West Bengal. More than 25 responses each came from Haryana, Madhya Pradesh, Chhattisgarh, Bihar and Gujarat.

**Table 11:** Safety perceptions during duty hours by region. shows that safety perception trends are comparable across regions in India, with minor differences that can be expected due to sample size from each region.

| Region | Very Unsafe | Felt Unsafe | Intermediate | Safe | Safest | Total |
| --- | --- | --- | --- | --- | --- | --- |
| **Central India** | 8 (11.9%) | 15 (22.4%) | 28 (41.8%) | 14 (20.9%) | 2 (3.0%) | 67 (100.0%) |
| **Eastern India** | 37 (18.0%) | 47 (22.9%) | 89 (43.4%) | 26 (12.7%) | 6 (2.9%) | 205 (100.0%) |
| **Islands and UTs** | 3 (8.1%) | 10 (27.0%) | 17 (45.9%) | 6 (16.2%) | 1 (2.7%) | 37 (100.0%) |
| **North-Eastern India** | 8 (12.3%) | 10 (15.4%) | 36 (55.4%) | 6 (9.2%) | 5 (7.7%) | 65 (100.0%) |
| **Northern India** | 40 (11.3%) | 95 (26.8%) | 173 (48.9%) | 38 (10.7%) | 8 (2.3%) | 354 (100.0%) |
| **Southern India** | 315 (11.4%) | 679 (24.6%) | 1258 (45.7%) | 388 (14.1%) | 115 (4.2%) | 2755 (100.0%) |
| **Western India** | 31 (7.7%) | 80 (19.9%) | 204 (50.7%) | 68 (16.9%) | 19 (4.7%) | 402 (100.0%) |
| **Total** | 442 (11.4%) | 936 (24.1%) | 1805 (46.5%) | 546 (14.1%) | 156 (4.0%) | 3885 (100.0%) |
| **Fishers Exact Test, p=0.024\*** |  |  |  |  |  |  |

**Table 12:** Safety perceptions during duty hours by work sector. shows subjective safety perceptions among healthcare workers in the government and private sector, revealing a significant difference between the two groups (Chi-square test, p < 0.0001).

| **Safety Perception** | **Government (N=792)** | **Private (N=724)** | **Total (N=1516)** |
| --- | --- | --- | --- |
| **Very Unsafe** | 135(17.05%) | 40(5.52%) | 175 (11.5%) |
| **Felt Unsafe** | 217(27.4%) | 87(12.02%) | 304 (20.1%) |
| **Uncertain Safety** | 356(44.95%) | 320(44.20%) | 676 (44.6%) |
| **Safe** | 69(8.71%) | 203(28.04%) | 272 (17.9%) |
| **Safest** | 15(1.89%) | 74(10.22%) | 89 (5.9%) |
| **Total** | 792 (52.2%) | 724 (47.8%) | 1516 (100.0%) |
| **Chi Square Test, p<0.0001 shows significance** |  |  |  |

**Table 13.** Suggestions for better workplace safety from individual doctors, ranked by frequency. *\*As the questionnaire directly mentioned duty room, ranking of this parameter is not possible. The other parameters were derived from individual comments and ranked according to frequency of use.*

|  |
| --- |
| 1. Duty room with bathroom, close to work location* |
| 2. Adequate number of professional security personnel |
| 3. CCTV camera 24/7 to monitor key areas |
| 4. CPA Central Protection Act, and its enforcement |
| 5. Secure locks/bolts on all duty room doors |
| 6. Restrict the number of bystanders/attendants with each patient |
| 7. Adequate lighting to be ensured |
| 8. Panic button/alarm system/code grey in case of emergency |
| 9. Food and water to be made available nearby during night duty |
| 10. Do not allow drunk bystanders in casualty |

The survey identified several rectifiable reasons for this.

The availability of duty room and facilities was limited, with only 55% being provided a duty room. Thus, 45% of doctors are compelled to find less secure places such as empty outpatient beds or seminar rooms to sit down or lie down. Nearly one-thirds of the available duty rooms did not have an attached bathroom, which means that the doctors needed to step outside during late hours to access these facilities.

Where available, duty rooms were often reported to be inadequate in size, safety, comfort, privacy, location and upkeep – all of which are factors that necessitate doctors to venture out to less secure places at night. Providing a safe place for doctors to rest during night duty is a basic necessity and does not come at great cost to any healthcare establishment. The survey unearths the lack of attention to this aspect of healthcare.

The data indicate that the presence of a duty room with an attached bathroom significantly improved the sense of safety. This correlation might be attributed to the possibility that hospitals with well-equipped duty rooms are also more likely to have invested in comprehensive security measures.

Apart from being a basic necessity, the availability and adequacy of these facilities translates into better patient care as doctors feel secure, safe and well-cared for.

Several additional factors were highlighted by doctors who participated in the survey. The lack of sufficient numbers of trained security personnel, inadequate lighting of the corridors, absence of CCTV cameras and unrestricted entry of unauthorised individuals into patient care areas were among the most frequent remarks. Providing basic facilities like clean and accessible bathrooms, drinking water and food for those who work through the night are expected of any healthcare establishment. When these are not provided, doctors are forced to walk through dark and insecure areas of the building or premises that are often not walled or fenced from the public road outside.

Some doctors indicated the need to start carrying weapons for self-defence. One doctor admitted that she always carried a foldable knife and pepper spray in her handbag because the duty room was located at the far end of a dark and deserted corridor. Doctors who worked in casualty reported verbal and physical threats from people who were drunk or under the influence of drugs. Another doctor reported that she repeatedly experienced bad touch or inappropriate contact in a crowded emergency room. The situation is worse in some smaller hospitals where there is limited staff and no security.

Several doctors reported apathy from the administrators when security concerns were raised, a common excuse being that the seniors also had endured similar working conditions. It is noteworthy that violence is predominantly experienced by junior doctors, who, being on the frontline, are particularly vulnerable but have limited involvement in administration or policymaking. Senior faculty members bear the responsibility of implementing policies to improve patient care delivery as well as enhancing security measures, thereby creating a safer work environment for junior doctors.

Doctors across the country have called for a Central Protection Law to prohibit violence in all healthcare settings and enforce airport-like security measures, ensuring a safer working environment and better patient care. Such a law would standardize security arrangements across the sector, ultimately benefiting patients as well as doctors.

The survey findings have significant implications for broad policy changes, some of which have already been addressed by the Government of India in response to the Kolkata incident. The Supreme Court of India took suo motu cognizance, assuring that “doctors and medical professionals shall stand assured that their concerns are receiving the highest attention from the highest court, with input from a diverse range of experts.” [9,10]. The multiple factors that contribute to healthcare violence in India have been analysed earlier [11].

The large number of respondents with representation from multiple states in India from both public and private sector, across a broad range of age and professional seniority are the strengths of the study. All studies have limitations. While the anonymity offered by the online survey encouraged doctors to be frank and open about their concerns, individual comments cannot be verified. A significant limitation of any survey is the potential for sampling bias. Participation may have been skewed towards doctors with strong opinions or personal experiences related to safety. However, the clear differences observed between public and private sectors, as well as by professional seniority, suggest that the survey captured genuine and relevant responses. The findings may be supplemented with in-person interviews by trained professionals.

## 7. Conclusion

Doctors across the country report feeling unsafe during night shifts. This survey, the largest of its kind in India, identifies several modifiable risk factors contributing to violence in healthcare settings. There is significant potential for enhancing security personnel and equipment. Infrastructure improvements are needed to ensure that duty rooms, bathrooms, food, and drinking water are safe, clean, and accessible. Adequate staffing, effective triaging, and crowd control are essential in patient care areas to ensure that doctors can provide necessary attention to each patient without feeling threatened. The extensive and diverse suggestions from individual doctors, summarised and quoted in this article, will provide valuable insights for administrators and policymakers.

## Data Availability

All data produced in the present study are available upon reasonable request to the authors but anonymity of people who contributed data will be maintained

## 9. Competing Interest Statement

The authors have declared no competing interest.

## 10. Funding Statement

No funding involved.

## 11. Author Declarations

We confirm all relevant ethical guidelines have been followed, and any necessary IRB and/or ethics committee approvals have been obtained.

## Notes

### Funding Statement

This study did not receive any funding

### Author Declarations

The Indian Medical Association Ethics committee approved this study. This included experts including Dr Srikumar Vasudevan, Dr VD Pradeepkumar, Dr Abraham Paul

